# Impact of Whole Slide Image Blurriness on the Robustness of Artificial Intelligence in Real World Setting: Retrospective Observational Study

**DOI:** 10.1101/2025.03.05.25323268

**Authors:** Ho Heon Kim, Young Sin Ko, Kyungeun Kim

## Abstract

**Context:** In digital pathology, blurriness in whole slide images (WSI) is a common issue, with severe blurriness widely acknowledge as a critical factor that can degrade the performance of artificial intelligence (AI) models. However, the effects of the typical levels of blurriness observed in real-world pathological images on the robustness of AI predictions remains unclear and unexplored.

**Objective:** To evaluate the impact of WSI blurring on the robustness of AI prediction in real-world setting.

**Design:** A retrospective study was conducted using 8,000 WSIs and corresponding AI predictions from four AI models trained on data from two scanners and two organs. WSIs were categorized into concordant and discordant groups based on AI-prediction accuracy. Analyses included: 1) comparing blur metrics between groups, 2) determining the odds ratio between the proportions of blurry patch in WSIs and prediction concordance, and 3) assessing model performance across varying blur intensities.

**Results:** For each organ-scanner pair, the average wavelet score and Laplacian variance for WSIs between the two groups did not show a statistically significant difference model (p > 0.05 for both metrics), except for one, and their effect sizes were small (Cohen’s *D* < 0.2 for both metrics). Additionally, no statistically significant association was observed between AI prediction concordance and the proportion of blurry images in WSIs (confidence intervals included 1, respectively). Model performance remained robust even at high blur level (radius=1) at which patch image had Laplacian variance of 162.88 and a wavelet score of 1880.07, corresponding to the top 1.22% and 2.16% of blurriness respective, in our dataset.

**Conclusions:** The findings empirically suggest that the typical levels of WSI blurriness encountered in real-world settings may not significantly compromise the robustness of AI predictions.

## Introduction

Recent advancements in computer hardware and algorithms have significantly enhanced the ability to process large-scale data, leading to the widespread adoption of digital pathology and AI application across many institutions^1^. From basic histopathological task such as tumor detection to the advanced task such as genotype prediction, and survival prediction, AI in digital pathology have demonstrated substantial clinical impact on the diagnostic process^1,2^. Empirical evidence from numerous studies indicates that AI techniques not only improve the accuracy of pathological diagnoses but also enhance diagnostic efficiency in clinical practice ^3–6^.

However, despite the increasing integration of digital pathology and AI into pathology workflows, concerns remain regarding the robustness of AI predictions. These concerns often arise from the quality or variability of WSIs, including image artifacts, and inconsistency in color during the digitalization process of slide images^7–9^. Among these artifacts, the most common artifact found in WSI is image blurriness, which is mainly caused by misalignment with focal plane of the scanner during digitialization^10^. This artifact is largely due to scanners, which cannot produce completely identical digital images, even when the same slide is scanned repeatedly with the same device^11^. Once WSIs are scanned, restoring severe out of focus (OOF) images to their original quality is challenging unless the slides are rescanned in the practice^12,13^. Because, in real-world clinical workflows, rescanning is often impractical due to time constraints and the additional workload it imposes on pathologists and laboratory staff.

To address these challenges, several studies have investigated methods for detecting blurry images using quantitative measurements, or AI-based techniques^11–17^. Building on this, subsequent research has proposed deep learning models for deblurring defocused pathology images^13,18^. However, there is a lack of evidence to substantiate the assertion that blurriness of image is significantly problematic in real-world scenarios. To our knowledge, only one study has examined the impact of image blurriness on machine learning prediction^11^. However, the generalizability of its finding remains limited due to methodological constraints. Specifically, the study employed conventional machine learning approach, such as support vector machines, rather than deep learning, which dominates contemporary pathology research. Moreover, the study was conducted using a small dataset with limited image specification (e.g. the number of sections per slide), and a narrow range of scanners and organs. These limitations make it challenging to generalize its conclusions or use them as evidence to support the application of deep learning in this context.

Therefore, a comprehensive investigation encompassing how blurriness affects AI predictions is needed particularly in terms of the proportion of blurry images within WSIs, the severity of the blur, and its variability across different scanners, organs, and AI models. The aim of this to determine whether WSI blurriness compromises the robustness of AI prediction models in digital pathology in real-world clinical settings.

## Method

### Slide preparation & Digitalization

Hematoxylin and eosin (H&E) slides for WSI were prepared according to the standard operating procedure. The specimen obtained by biopsy, polypectomy and endoscopic resection were fixed in 10% neutrally buffered formalin and embedded for paraffin blocks. Paraffin blocks were sectioned to 3 μm thickness, and at least three consecutive sections were mounted on one slide. H&E staining was performed using an autostainer (Dako Cover stainer2 CS100, Agilent). The prepared slides were randomly assigned to different scanners for digitization.

### SeeDP: Diagnostic quality control system

As one of the clinical reference laboratories, the Seegene Medical Foundation has developed an AI-driven quality control system called SeeDP^5^. SeeDP consists of several components: 1) a backend system that interacts with the laboratory information system; 2) a web interface for visualizing AI prediction results; and 3) an AI system that classifies WSIs that have already been pathologically diagnosed. The AI system consists of separate AI models for each organ and scanner. The AI model itself is composed of two submodels: a patch-level model and slide-level model. The patch-level model predicts lesion classes at the patch level, while the slide-level model aggregates the patch-level predictions to generate a final slide-level classification. Specifically, four AI models have been trained, one for each combination of the two scanners (Leica Aperio GT450, hereinafter referred to as GT450, and 3DHISTECH PANNORAMIC 250 Flash III, hereinafter referred to as P250) and two organs (stomach, hereinafter referred to as S, and colon, hereinafter referred to as C). These four combinations are collectively referred to as organ-scanner pairs throughout this study. Unless otherwise specified, statistics are presented in the order of S-GT450, S-P250, C-GT450, and C-P250.

Once a WSI is generated, the backend system detects the newly created slide and sends a request to the AI system to predict its classification into one of three categories: 1) normal, 2) dysplasia, and 3) malignant, the detailed criteria for categories are as described in a previous paper ^6^. Upon completion of the AI prediction, pathologists review any discrepancies between their pathological conclusions and the AI’s predictions. If necessary, they can revisit and reassess their original interpretations using the web interface^5,6^.

### Study design & Sample selection

We retrospectively collected pathology reports of colon and stomach specimens and their WSIs after pathology diagnosis on SeeDP system (September 1, 2023 to September 1, 2024). A total 265,557 WSIs from 223,629 patients were initially collected, and among them, 4,162 WSIs (1.57%) were excluded due to scanning errors, such as extremely small size of specimen on slide, or absent tissue on the slides or severe OOF that the scanner automatically flagged as error. The WSIs classified as errors in the scanner were not scanned again and were ultimately not included in the SeeDP AI prediction.

For the remaining 261,395 WSIs, we grouped them based on organ and scanner (Figure 1). For each pair in the scanner-organ pairs, we partitioned the data based on the concordance between pathologists’ diagnoses and AI predictions to identify the association between image blur and concordance. This study design is analogous to a basic epidemiological approach, such as a “case-control study,” which compares cases with the outcome to those without the outcome to investigate the association between exposure and the frequency of the outcome^19^. We randomly sampled 1,000 WSIs from each subgroup due to computational cost. In total, 8,000 WSIs were analyzed (Figure 1).

**Figure 1.**
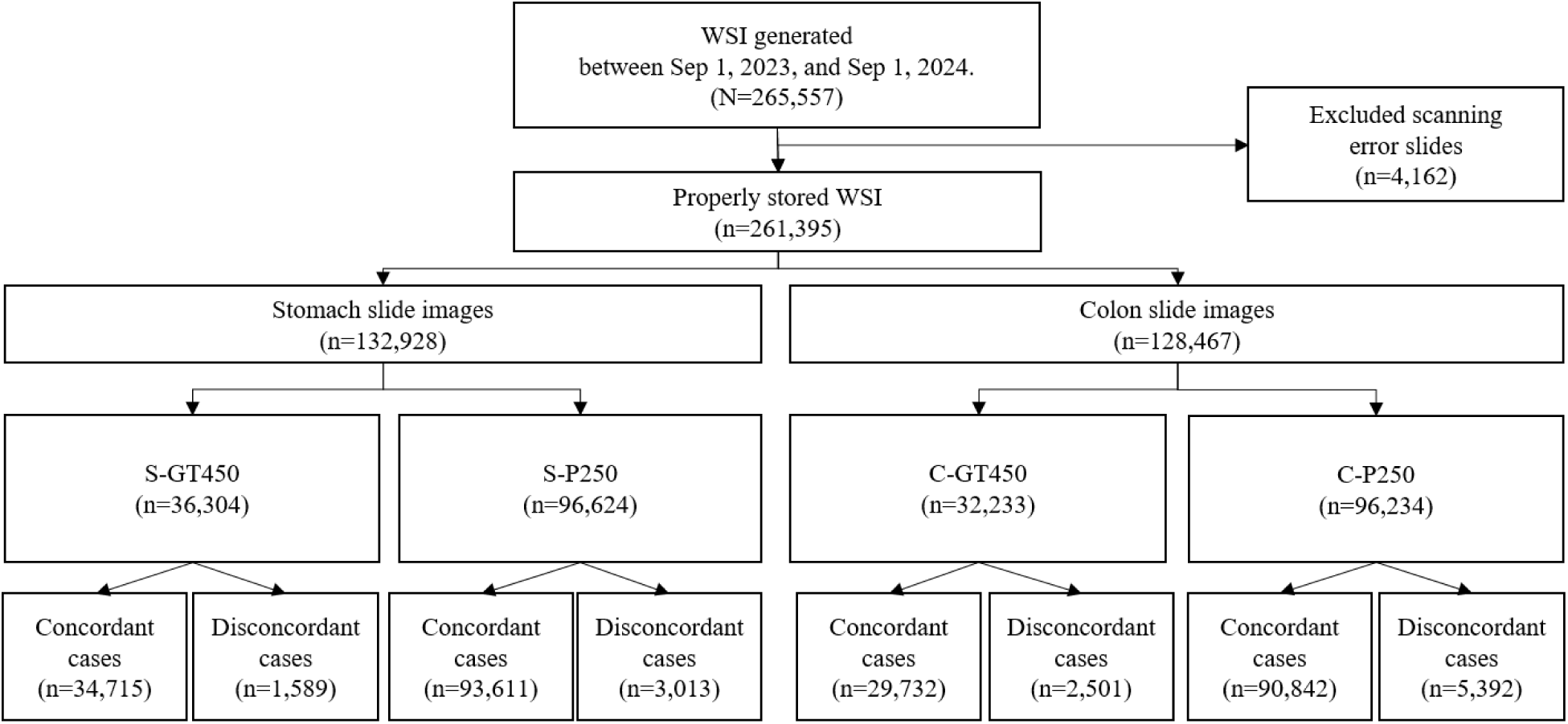
Selection flow of eligible WSI

### Image preprocessing

WSIs, generated from slide scans, are large-scale images, reaching up to approximately 300,000 pixels depending on the scanner. It is hard to directly process the large-scale image once on CPU memory. We performed tessellation by dividing each WSI into multiple 256 x 256 patches. To remove background patches, we employed JPEG compression. After the compression, the image file size was calculated to estimate the proportion of non-background areas. Since JPEG compression efficiently encodes redundant low-frequency regions, such as backgrounds, it achieves a higher compression rate for non-tissue areas, resulting in smaller file sizes. Consequently, patches with a JPEG file size below a predefined threshold were excluded (10 KB), as they primarily represented background regions. This approach ensured that only relevant tissue regions were retained for further analysis, improving the model’s training efficiency and accuracy. To handle the computational workload efficiently, we utilized a clustered computing service with job scheduling tools (Slurm Workload Manager) across 4 machines, each equipped with 128 CPU cores.

### Quantitative measurement of blurry image

To quantify image blur, we employed two metrics. The first was Laplacian variance, a widely used spatial-domain method for edge detection. This method measures local pixel intensity variation, effectively capturing regions with rapid intensity changes, which typically correspond to edge and in-focus areas^20^. In this approach, the Laplacian operator, a second derivative operator, identifies areas of rapid pixel intensity changes, with blurry images exhibiting weaker or fewer edges, resulting in lower Laplacian variance values^21^. To implement this, we first converted RGB patch into gray patch. Then, we convolved the gray patch with 3 x 3 positive Laplacian kernel to detect edges intensity. Finally, we calculated the variance of the edge intensity, as the variance reflects the spread or variability of the edge intensity values. The lower Laplacian variance indicates a blurrier image.

The second metric was “wavelet score” was based on spectral-domain methods, which analyze frequency components of an image^20^. High-frequency components correspond to sharp regions, while low-frequency components are associated with blurred region. Specifically, we employed the Haar Wavelet Transform (HWT) and calculated the sum of the high-frequency sub-bands (e.g. high-high, high-low, and low-high) from the HWT decomposition. This summed statistics serves as measure of the high-frequency content reflects image sharpness or complexity of image^22^. Similar to Laplacian variance, a lower wavelet score indicates a blurrier image.

### Statistical analysis

To figure out whether the quality of WSI blurring affects the robustness of AI prediction models, we conducted three-part analysis. First was the comparative test which compared the blur-related statistics of concordantly predicted WSIs and that of discordantly predicted WSIs. Since WSIs were divided into multiple patches, representative statistics (minimum, maximum, and average of blur-related metrics) were calculated to summarize slide-level characteristics. We assumed that the sharpness metrics of slides with discordant predictions would be statistically lower than those of slides with concordant predictions.

Secondly, the proportion of blurry patches within WSIs was evaluated, with the hypothesis that a higher proportion of blurry patches would be associated with reduced prediction accuracy. For this analysis, blurry patches were defined as those with blur metric values in the bottom 5% of all patch images, which is similar with previous study^11^. To calculate the proportion of blurry images, we first identified the number of blurry images according to the definition of blurry patch among the foreground patches for a WSI, and then computed the proportion of blurry patches by dividing this count by the total number of foreground patches. We then fitted a logistic regression model to estimate the coefficient for the proportion of blurry patches in relation to the discordance in AI predictions.

Thirdly, to assess the robustness of our model against blur, we employed artificial blur, as it is not feasible to replicate out-of-focus conditions in already scanned images. Gaussian blur was applied to patches, with varying kernel sizes to control the intensity of the blur. We evaluated the model’s predictive performance, specifically AUROC, under these conditions to determine the threshold at which blur caused significant degradation in predictions. Concordantly predicted cases on high-quality images served as the reference. This allowed us to measure the impact of artificial blur on model performance across varying levels of image degradation.

For statistical analysis, we performed independent t-test with *p*-values to assess the statistical significance of the differences of continuous variables between the two groups. Additionally, we conducted a chi-square test to evaluate the statistical significance of differences in proportions across categories between the two groups. A *p*-value threshold of 0.05 was used to determine statistical significance. Furthermore, effect sizes were calculated to assess the magnitude of differences, providing additional context beyond statistical significance. As the statistical significance was not adequate enough to compare groups of a large population, which will almost always demonstrate a significant difference because of statistical power. To measure the magnitude of difference in the mean of two groups, Cohen’s d was calculated. Cohen’s *d* is considered as small (0.2≤d<0.4), medium (0.4≤d<0.8), and large (d ≥ 0.8)^23^. For categorical variables, Cramer’s V was calculated and interpreted as follows: negligible (0.0≤V< 0.1), weak (0.1 ≤ V < 0.2), moderate (0.2 ≤ V < 0.4), strong (0.4 ≤ V).

### Ethics

The study was approved by the institutional review board at Seegene Medical Foundation (SMF-IRB-2024-015). The anonymous and deidentified nature of the retrospective pathologic image made obtaining informed consent unnecessary.

## Results

### WSI characteristics

From a random sample of 8,000 WSIs, we obtained a total of 3,433,212 patches. For stomach samples, most of the pathologists’ diagnoses were classified as gastritis (N class according to our categories) (S-GT450: n=1,601, 80.05%; S-P250: n=1,477, 73.85%). Similarly, for colon samples, the majority of pathologists’ diagnoses were normal for the C-P250, while dysplasia was predominantly diagnosed for the C-GT450 (Table 1). Among the normal stomach samples confirmed by pathologists, 628 (15.70%) of the WSIs were incorrectly predicted as dysplasia across both scanners. For normal colon WSIs, 812 (20.30%) were incorrectly predicted as dysplasia (Supplementary figure 1). Although differences in the proportion of pathologists’ diagnoses between GT450 and P250 were observed in both organs, the magnitudes of these differences were negligible and weak (Cramer’s V = 0.096 and 0.119, respectively).

**Table 1.** Characteristics of WSIs. Top: WSI of the stomach; Bottom: WSI of the colon.

| Variables | Stomach-scanner pairs<br>(n=4,000) |  | *P* value | Effect size<br>(Cramer's *V*) |
| --- | --- | --- | --- | --- |
|  | S-GT450<br>(n=2,000) | S-P250<br>(n=2,000) |  |  |
| Pathologist's diagnosis (n, (%)) |  |  | <.001 | 0.096 |
| Normal | 1,601 (80.05) | 1,477 (73.85) |  |  |
| Dysplasia | 221 (11.05) | 221 (11.05) |  |  |
| Malignant | 178 (8.90) | 302 (15.10) |  |  |
| AI prediction(n, (%)) |  |  | <.001 | 0.203 |
| Normal | 1,213 (60.65) | 1,198 (59.90) |  |  |
| Dysplasia | 321 (16.05) | 584 (29.20) |  |  |
| Malignant | 466 (23.30) | 218 (10.90) |  |  |

| Variables | Colon-scanner pairs<br>(n=4,000) |  | *P* value | Effect size<br>(Cramer's *V*) |
| --- | --- | --- | --- | --- |
|  | C-GT450<br>(n=2,000) | C-P250<br>(n=2,000) |  |  |
| Pathologist's diagnosis (n, (%)) |  |  | <.001 | 0.119 |
| Normal | 904 (45.20) | 1,026 (51.30) |  |  |
| Dysplasia | 1,065 (53.25) | 880 (44.00) |  |  |
| Malignant | 31 (1.55) | 94 (4.70) |  |  |
| AI prediction(n, (%)) |  |  | <.001 | 0.094 |
| Normal | 967 (48.35) | 855 (42.75) |  |  |
| Dysplasia | 882 (44.10) | 885 (44.25) |  |  |
| Malignant | 151 (7.55) | 260 (13.00) |  |  |

### Quantitative assessment of blurriness

To summarize the characteristics of multiple patches within a single WSI, representative statistics were calculated as summary metrics for each WSI. Among these, the minimal values of Laplacian variance in concordant slides did not consistently higher than those in discordant slides across all scanner-organ pairs suggesting that the most blurred regions in WSIs with discordant predictions did not always appear noticeably blur than those in WSIs with concordant predictions (only for S-P250, C-GT450, p<0.05 and Cohen’s D=0.23; 0.21) (Figure 2A). In contrast, although the statistical significance varied across organ-scanner pairs, discordant WSIs consistently exhibited higher average and maximal Laplacian variance than concordant WSIs (Figure 2). These findings suggest sharpness of WSI with discordant prediction did not consistently show blur than that of WSIs with concordant prediction.

**Figure 2.**
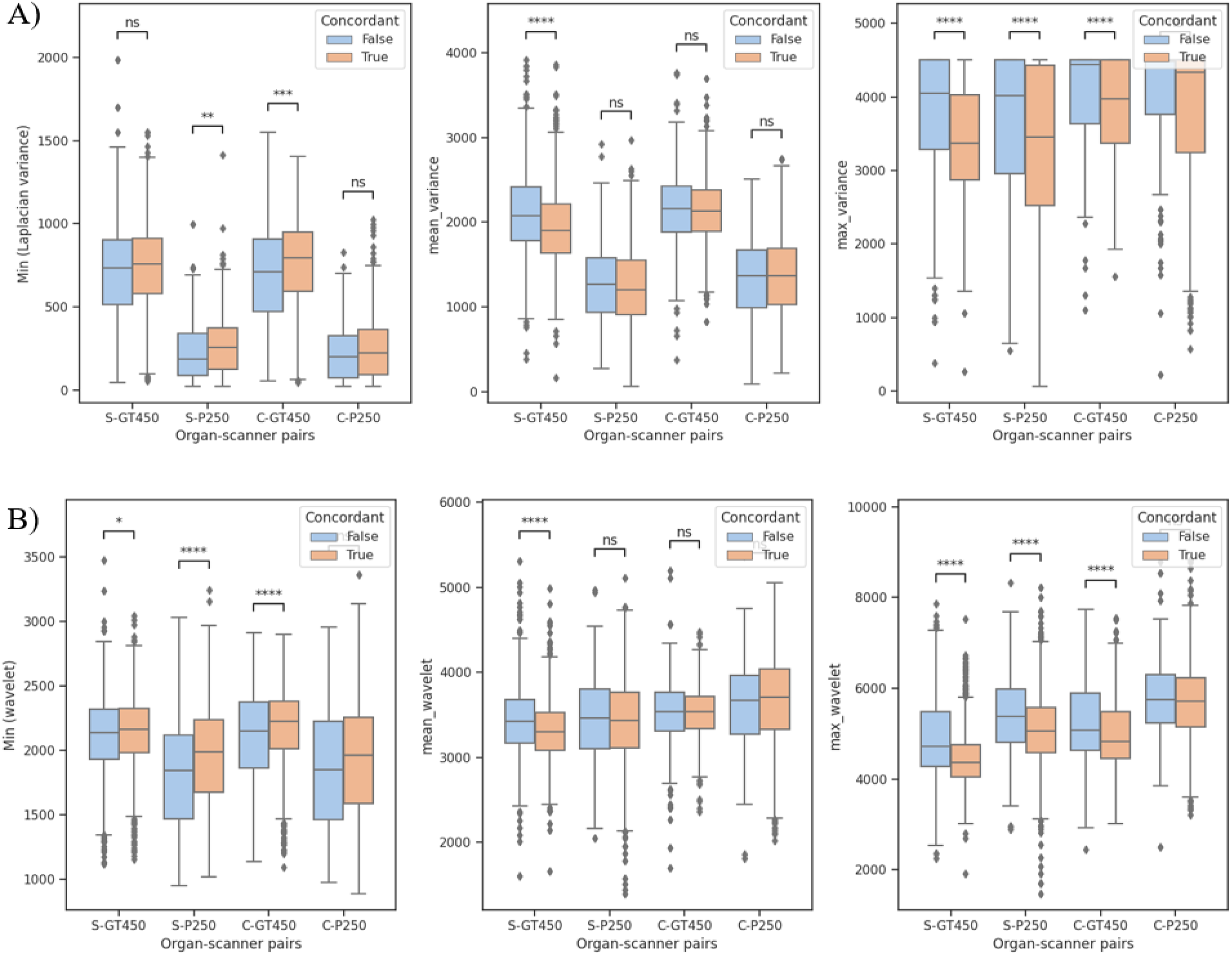
Blurry Image Assessment Using Representative Statistics for WSIs. (A) Minimum, average, and maximum Laplacian variance for each group. (B) Minimum, average, and maximum wavelet score for each group.

For the wavelet score, representative statistics showed trends consistent with those observed for Laplacian variance: (1) The minimum wavelet score in concordant slides did not exhibit a moderate or greater difference compared to discordant slides (Cohen’s D=0.23; 0.21, 0.07 for S-P250, C-GT450, C-P250 respectively), (2) the average wavelet score did not show consistent difference, and (3) the maximum wavelet score was also lower in concordant slides (Figure 2-B). Across both metrics, no substantial blurriness differences were observed between discordant and concordant WSIs, implying that image sharpness is not associated with diagnostic concordance.

### Association blur image ratio and AI prediction concordance

Logistic regression analyses were conducted to assess the association between the proportion of blurry patches in a WSI and AI prediction concordance (Figure 3). For Laplacian variance, all 95% confidence intervals (CIs) included 1, indicating no significant association. Similarly, for the proportion of blurry patches measured using wavelet scores, the 95% CIs for all regression coefficients also included 1. With both measurements, a higher proportion of blurry patches did not correspond to an increase in AI prediction discordance (Figure 3).

**Figure 3.**
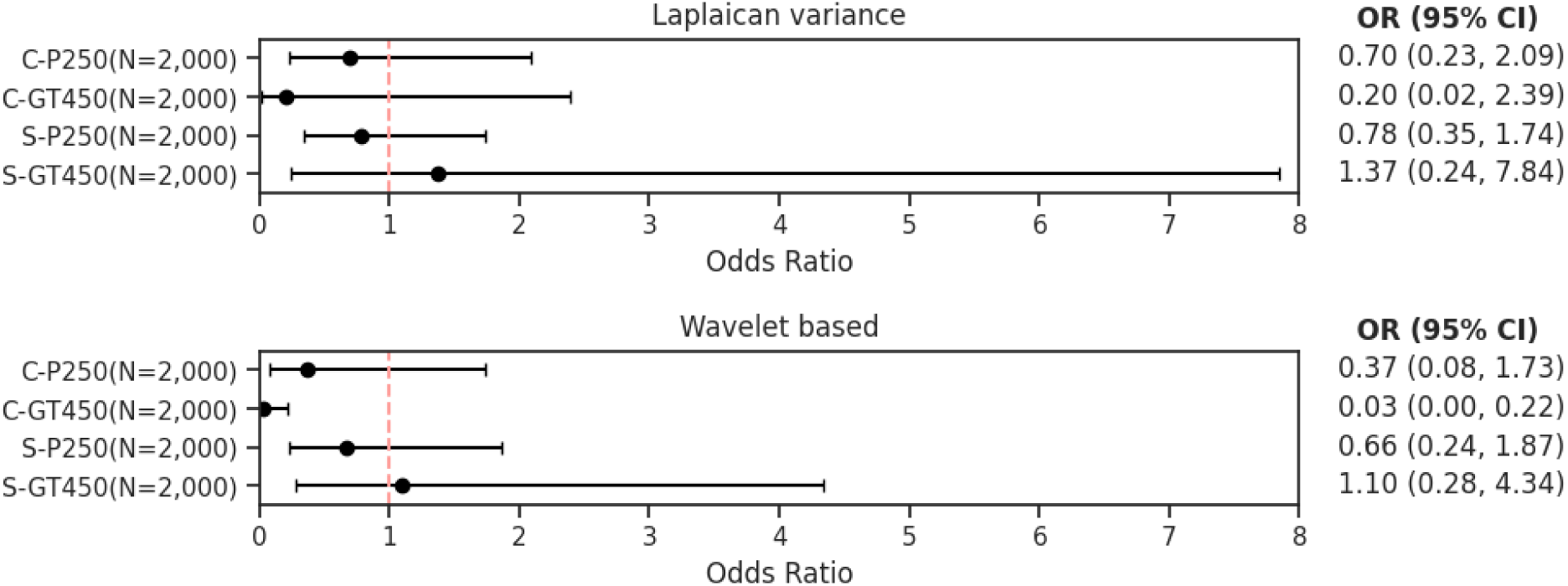
Forest plot showing the association blur image ratio in WSI and AI prediction concordance

After adjusting for a potential confounder, such as the pathological diagnosis of the slides, which may influence the concordance of AI predictions depending on the interpretative difficulty of the slides, the confidence intervals for the coefficients in all models consistently included 1. It suggests that a high proportion of blurry patches was not associated with AI prediction discordancy, even when accounting for pathological diagnosis (Supplementary figure 2).

### Blur intensity and AI prediction concordance

By applying artificial blur to the original patch images, our model maintained AUROC performance with slight decreases at a blur intensity of 1.0 (Radius=1) (Figure 4 and 5). At this intensity of artificial blur, the AUROC values for the four models decreased slightly from 1.0 to 0.966, 0.979, 0.990, and 0.996 for the three categories in the organ-scanner pairs respectively (Figure 4-A). The average Laplacian variance at a blur intensity of 1.0 was 157.07, with a standard deviation of 85.57 (1,828.23 [496.73] for wavelet) (Figure 4-B). This intensity represents a rare occurrences of severe blur image quality, as it corresponds to the 1.16th percentile for Laplacian variance (1.83th percentile for wavelet) (Figure 4-C). However, even at this level, the tissue structures within patches at 1.00 MPP remained sufficiently clear for distinguishing and identifying different tissue types (Figure 5).

**Figure 4.**
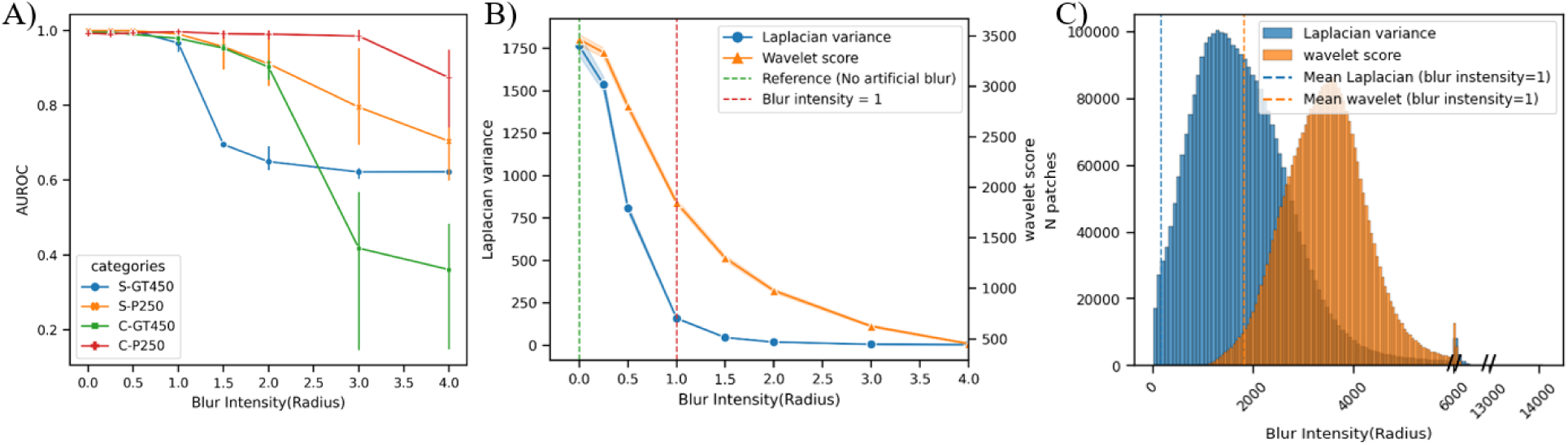
Blur intensity and AI prediction performance. A) average AUROC values across varying levels of artificially induced blur intensity for each subgroup; B) the blur intensity and blurry image metrics. The gray dashed line indicates a blur intensity level of 1. C) Distribution of blur metrics across patches. The dashed lines represent the average sharpness metrics at blur intensity level of 1.

**Figure 5.**
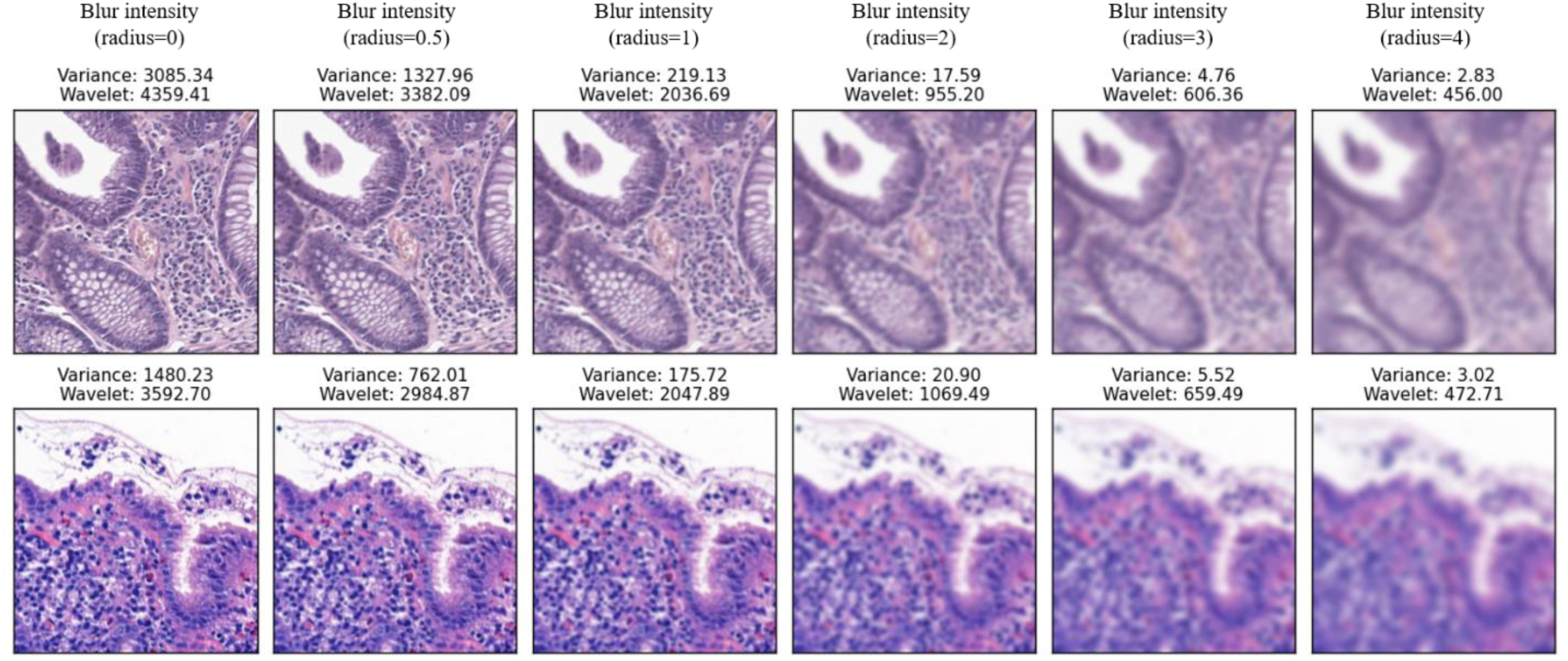
Example patch images by artificial blurring intensity

### Case review of achieving diagnostic concordance despite of blurriness

Colon biopsy and stomach biopsy cases diagnosed as tubular adenoma, low grade dysplasia by pathologists were well classified as dysplasia by SeeDP prediction, although significant blurriness was present ^6^ (Figure 6). Quantitative analysis of the WSI of colon adenoma revealed *regional* OOF patch images with high values of Laplacian variance (minimum, 102.62; mean, 1012.70; maximum, 4044.99) and 36.12% blurry patches according to our definition of blurry patches (Figure 6-A,B,C). Another gastric dysplasia case displaying intermediate grade blur with *global* OOF regions (76.1% of patches by the definition) and low values of Laplacian variance (minimum, 68.25; mean, 68.25; maximum, 358.44). The blur areas were located in third section in both cases, the SeeDP AI model consistently were consistently classified as dysplasia with blue color (Figure 6-A,E). In both cases, some histological characteristics of tumors, such as nuclear size and chromatin intensity, were preserved in the degree of blur that usually occur.

**Figure 6.**
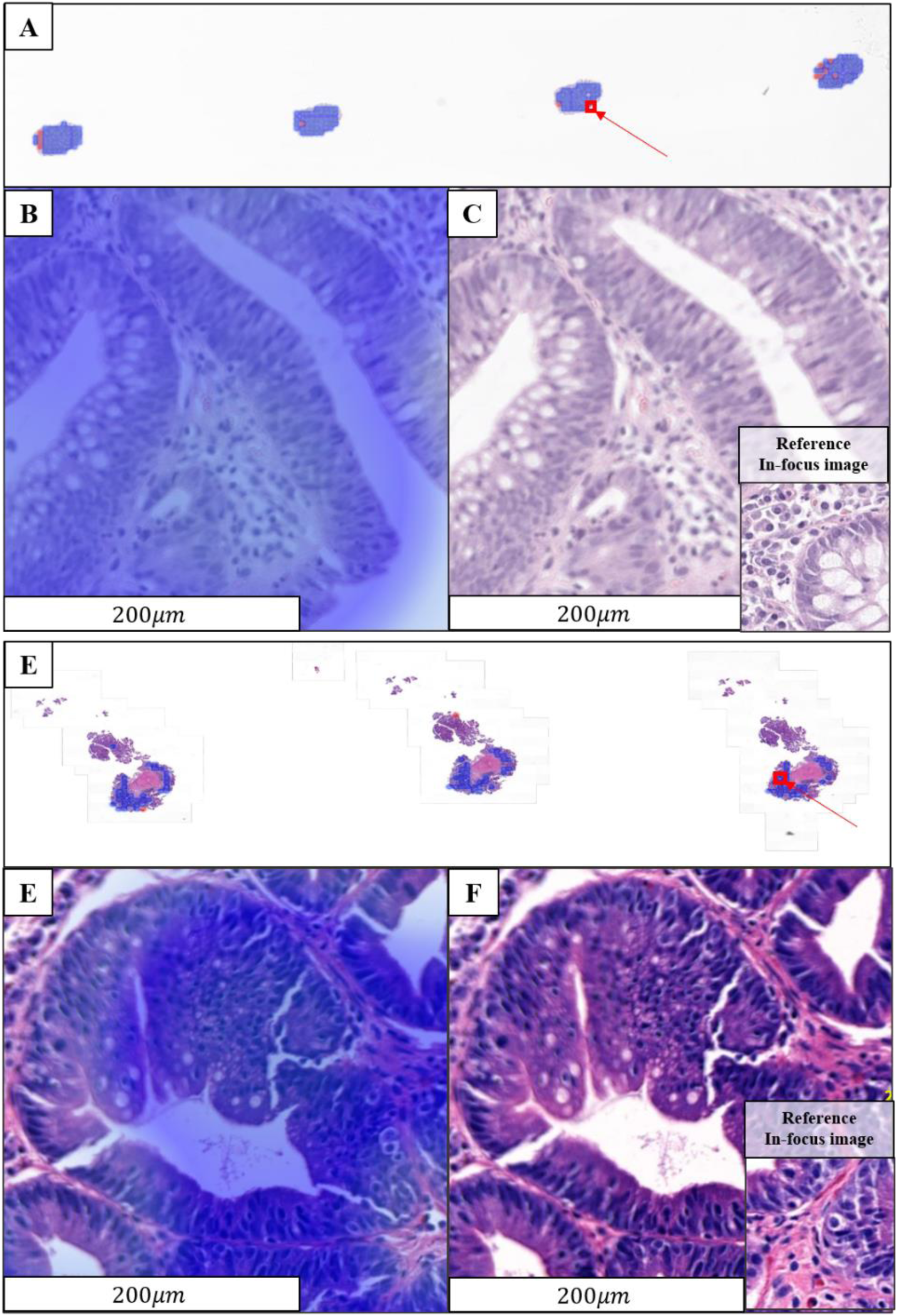
WSI of tubular adenoma in colon scanned with GT450 (C-GT450), and another WSI of adenoma scanned with P250 (S-P250). A) The red circles indicate AI-predicted malignant patches, and the blue circles indicate predicted dysplasia patches. Also, red box and arrow indicate the region of B, C. B) AI prediction in the lesion, C) original image in same lesion with B. D) The thumbnail of WSI of adenoma scanned with P250 (S-P250). E) AI prediction in the region of interest lesion in D. F) original image in same lesion with E.

## Discussion

### Principal findings

Many studies have proposed AI methodologies to address challenges posed by suboptimal image quality, particularly blurriness, to enhance the robustness of AI applications. However, in practical settings, the necessity of explicitly accounting for low-quality images during AI development remains an open question. To our knowledge, this is the first study to examine the relationship between blurry whole-slide images and AI predictions using deep learning in real-world scenarios. Our findings indicate that commonly observed blurry images do not significantly affect the performance of AI models. This conclusion is supported by experiments conducted across multiple scanners and models, including 4 organ-scanner pairs. Additionally, leveraging large-scale data from a reference laboratory in South Korea, our study utilized up to 8,000 slides to ensure the generalizability of these findings.

Additionally, our research demonstrated that the proportion of blurry patches was not correlated with AI prediction performance, contrary to expectations. We initially hypothesized that a higher proportion of blurry images would lead to an increased discordance rate between pathologic conclusions and AI predictions. However, statistical analysis did not confirm that a greater proportion of blurry images resulted in discordant predictions across four organs-scanner pairs (Figure 3). Notably, our observations revealed that the proportion of blurriness severe enough to adversely impact AI performance was rare, occurring in approximately 1% of the total patch images (Figure 4).

### Characteristics of WSI and AI model for robust predictions

We interpreted these findings on the robustness of AI predictions as being influence by two key factors: data-driven factors, which related to the unique characteristics of WSIs that are not typically encountered in natural images, and AI model-driven factors, which pertain to the robustness of the AI model itself.

Firstly, these data-driven factors arise from the unique distinctive nature of WSIs, which differ significantly from conventional natural images. In conventional image classification task using deep learning, models typically make predictions on small, individual images, such those in CIFAR-10 (32×32 pixels), and ImageNet (commonly resized into 256×256 pixels)^24^. In contrast, AI models for histopathology classify WSIs, which are composed of numerous patch images due to their gigapixel size. This patch-based composition allows AI predictions to remain robust at the WSI level, even when some patch-level predictions were misclassification due to patch blurriness.

In detail, OOF can be categories by three types: *global* OOF that affect the entire slides, *regional* OOF that affects large tissue patches, and *local* OOF that affect individual cells or subcellular structures^25^. In our observations, global OOF was an extremely rare event. In our observation, 98% of our WSIs had a blurry patch proportion of less than 0.51 based on Laplacian variance and less than 0.363 based on the wavelet score among 8,000 WSIs (Supplementary figure 3). Since deep learning models predict scores at the slide level, *regional* or *local* OOF would not critically impact slide-level AI predictions. During real-world AI deployment, we observed that the AI model could accurately predict malignancy for WSI with advance gastric carcinoma, even when large regional of the slide exhibited *regional* OOF (Supplementary figure 4). Particularly for biopsy slides, the lesions were sufficiently large, allowing the model to overcome potential prediction failures caused by blurriness through analysis of other regions. In addition, for the specimen with endoscopy resection, it is possible that a critical factor is that our WSIs contain multiple sections on a single. Even if a *regional* OOF occurs within a lesion in one tissue section due to the scanner, the other two tissue sections would be enough to correctly slide-level prediction.

Secondly, AI model-driven factors refer to model’s inherent ability to handle blurry images, with trainable parameters that enable the AI model perform well even when exposed to blurry images. In our study, four models were fine-tuned to histopathology images, based on pre-trained models using transfer learning. During the training phase, our model would learn the histopathological features even from blurry image because we did not intend to exclude blurry WSIs. This blurry image exposure could enable our networks to have resistance for blur images.

In the field of machine learning, the hypothesis that exposure to blurred images might be functionally beneficial for the visual system in image recognition has been tested in several studies^26–29^. These studies showed that exposure to blurred images enables convolutional neural networks to achieve robust performance during the training phase. Additionally, data augmentation by generating synthetic blurry images is commonly used to increase resistance to blur^30^. In an experiment, the model trained only on sharp images showed more deteriorated performance than the model exposed to the mixture of dataset composed of blurry image and shape images^26^. Considering previous evidence, our model would have resistance to blurry images due to the exposure of blurry images commonly observed in real-world scenarios.

### Limitation

Our study inherently has several limitations. Firstly, although we analyzed four models across two scanners and two organs, our study was conducted based on a single-center study. Since our observations were generated in a single reference laboratory, the findings may be biased, limiting its generalizability. However, given the lack of evidence addressing the impact of blurry images on AI performance in real-world scenarios, our findings are important as they lay the foundation for future research in this area. Secondly, our finding can be interpreted as limited because the significant blurry images would be excluded. In the process of selection flow, we excluded the scanning error WSIs, which may include the WSI with global OOF. The exclusion rates for each dataset were as follows: 0.10% (38/36,304), 2.26% (2,187/96,624), 0.08% (27/32,233), 1.98% (1,910/96,234), respectively, following the order of organ-scanner pairs. However, although these exclusion rates vary by scanner, they remain relatively low and fall within the range commonly encountered in real-world practice. Therefore, except for extreme cases that were automatically excluded and flagged by the scanner in practice, our study still supports the robustness of the AI model to blurry images.

## Conclusion

Typically observable blurry WSI would not affect slide-level AI predictions using deep learning in real-world scenarios.

## Data Availability

The data used in this study are not publicly available due to privacy and confidentiality restrictions

