## Supplementary figure for "Impact of Whole Slide Image Blurriness on the Robustness of Artificial Intelligence in Real World Setting: Retrospective Observational Study"

### Supplementary data 1

**Supplementary figure 1**. Sankey diagram from pathological conclusion and AI predicted conclusion

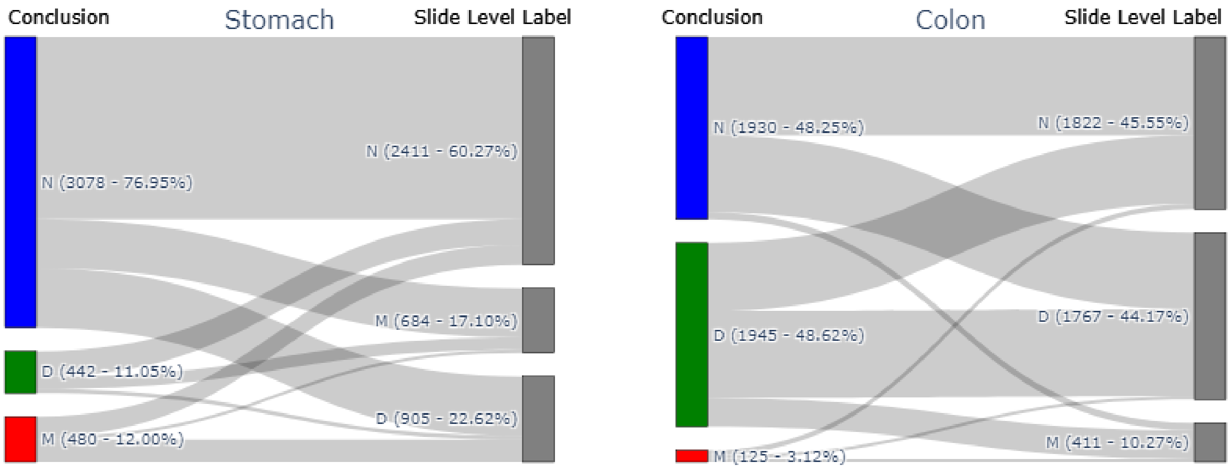

### Supplementary data 2

We additionally fitted a logistic regression model to adjust for confounding variables, such as class membership, which may influence prediction difficulty. For instance, discordant WSI predictions could be more attributable to the dysplasia class—known to be challenging for AI models to predict—rather than to image blurriness. To avoid multicollinearity arising from linear dependencies among class membership categories (e.g., normal, dysplasia, and malignant classes), we encoded binary variables for the normal and dysplasia classes only.

**Supplementary figure 2**. Forest plot showing the association blur image ratio in WSI and AI prediction concordance with the adjustment pathologic conclusion

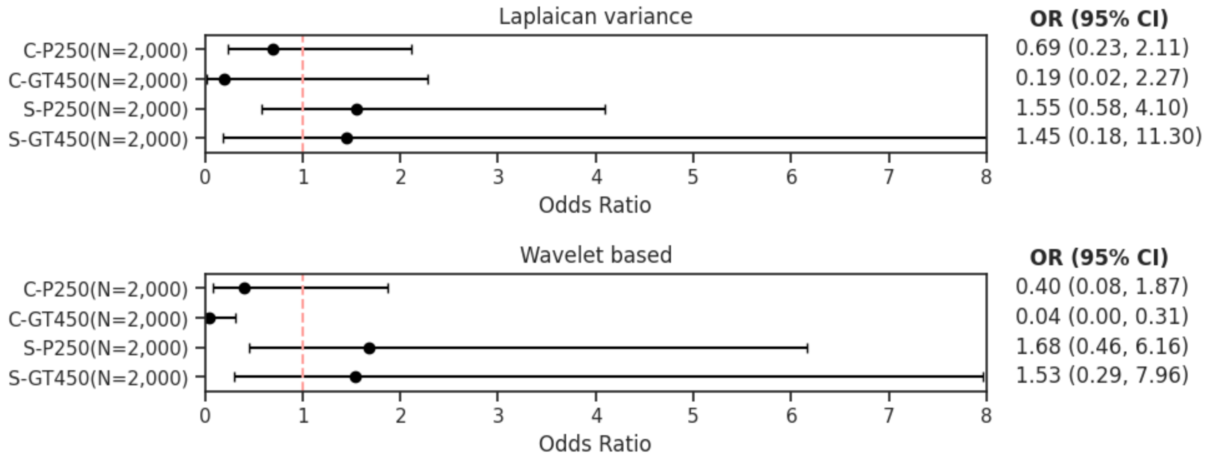

### Supplementary data 3

**Supplementary figure 3.** The proportion of blurry patches in a WSI

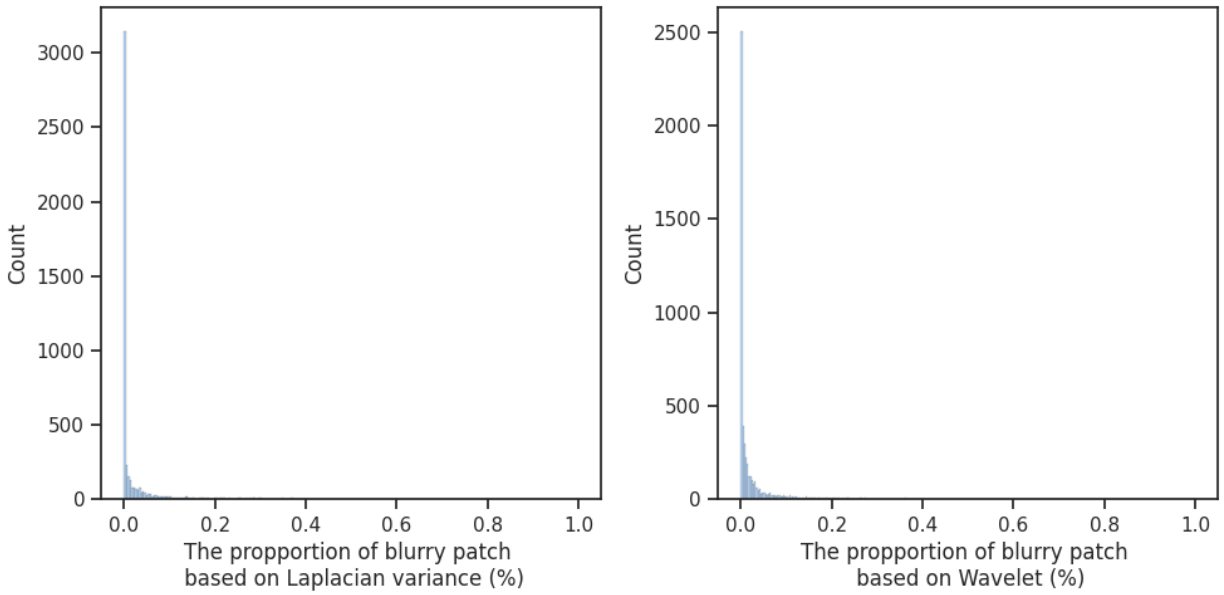

### Supplementary data 4

**Supplementary figure 4.** Regional out-of-focus whole slide image with advanced gastric carcinoma. A) Thumbnail of the whole slide image. The red circles indicate AI-predicted malignant patches, and the blue circles indicate predicted dysplasia patches. B) The region in the red box of subplot A

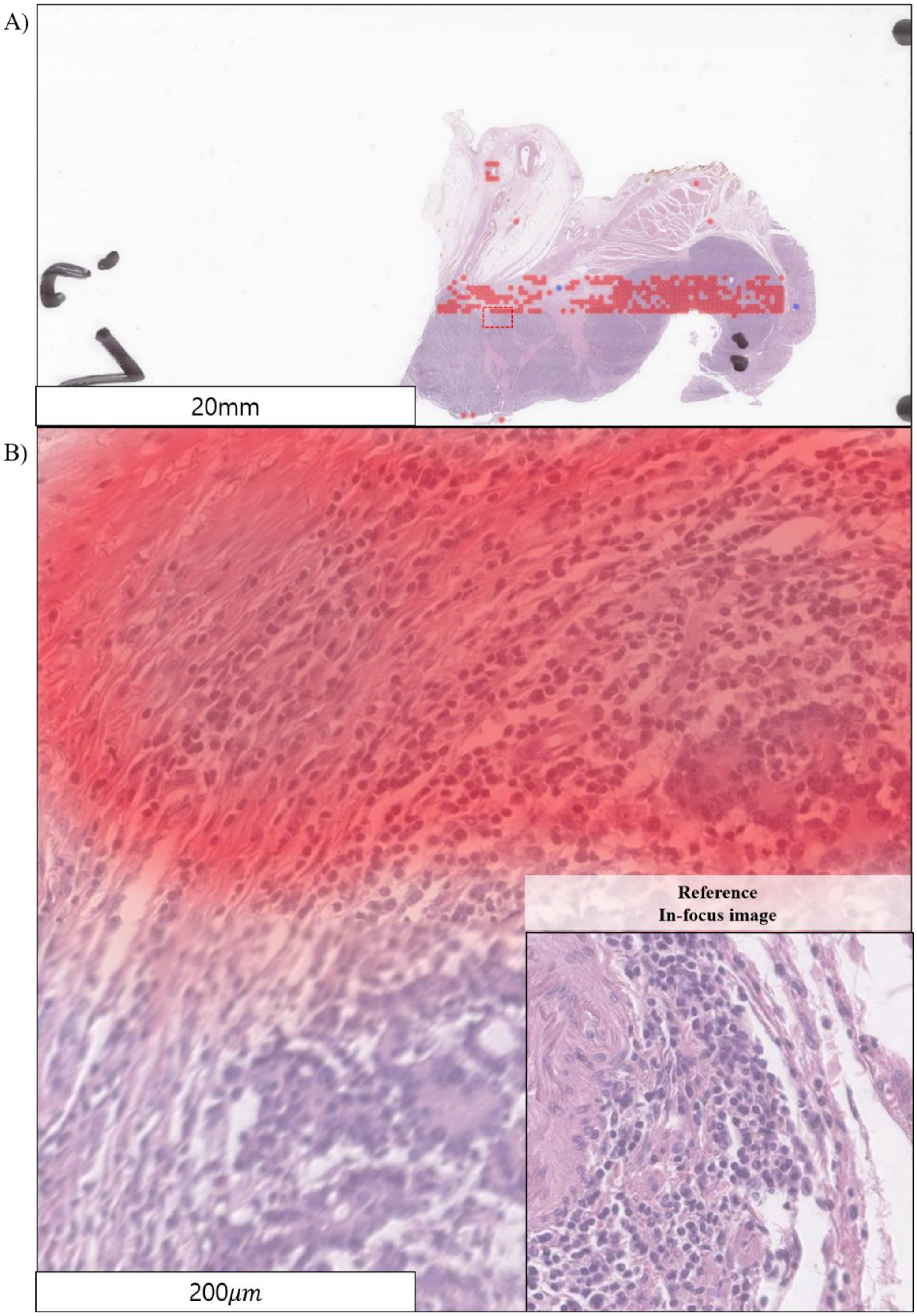
